# The experience of conducting non-pandemic research during the COVID-19 pandemic: a case study

**DOI:** 10.1101/2025.08.15.25333770

**Authors:** Micah J. June, Mary I. Otiti, Nicodemus O. Mbanda, Polland O. Miruye, Stephen J. Allen, Dónal O’Mathúna

## Abstract

**Background:** The COVID-19 pandemic presented unprecedented challenges to the conduct of non-pandemic research. The pandemic disrupted regulatory processes, strained health systems, and necessitated ethical recalibrations. This paper reports a retrospective study of the experience of conducting one clinical study of a non-pandemic related trial in western Kenya, conducted between October 2020 and January 2024. The original study documents were reviewed to identify lessons learned, particularly those relevant to ethical and regulatory aspects of conducting non-pandemic research during a pandemic.

**Methods:** A retrospective documentary case analysis was conducted using archived ethics and regulatory submissions, study progress reports, meeting minutes, and relevant peer-reviewed literature. A framework analysis approach guided thematic coding, supported by independent dual review to ensure reliability (Cohen’s Kappa ≥0.6).

**Results:** Public health restrictions during the third pandemic wave, particularly at the main recruitment site, compounded delays. Adaptive measures included reduced physical contact, use of satellite recruitment centers, community engagement with Traditional Birth Attendants, and remote monitoring via digital platforms. Parallel submission to both the Ethics Review Committee and the Regulatory Authority notably expedited the overall turnaround time for approval of protocol amendments. Despite these challenges, the study achieved an impressive 91% follow-up completion rate, with 96% of scheduled home visits successfully carried out.

**Conclusion:** This case study highlights how flexible, community-informed, and ethically responsive research practices can sustain clinical trials under adverse conditions. Strong stakeholder collaboration, open communication, and proactive risk mitigation were essential to maintaining study integrity and protecting participant welfare. These findings underscore the need for future research designs to embed contingency planning, community partnerships, and regulatory adaptability to ensure continuity and resilience for research conducted during public health emergencies.

## Introduction

Supportive research environments are critical for enabling high-quality, impactful studies while maintaining ethical integrity and fostering professional growth. The onset of the COVID-19 pandemic posed significant challenges to the global research ecosystem, particularly for non-pandemic-related research [1]. Ethical research conduct guided by principlism, including respect for autonomy, beneficence, non-maleficence, and justice [2] became more complex in a context marked by public health emergencies, shifting vulnerabilities, and constrained resources. In some settings COVID-19 presented a humanitarian situation requiring additional considerations for undertaking research that included examining the need to promote the health of communities, respect their dignity and uphold their rights [3]. Conducting non- pandemic research during a pandemic raises important questions about the prioritization of resources, public health responsibilities, and the distribution of benefits and burdens across society [4].

The pandemic required a realignment of research priorities and careful trade-offs in decision- making. The World Health Organization [5] emphasized the need to reassess ongoing and planned research projects, urging researchers and regulatory bodies to consider whether the risks posed by study participation in a pandemic context were ethically justifiable. Consequently, non-COVID-19 research was subjected to heightened scrutiny, new regulatory frameworks, and logistical challenges ranging from delays in ethical approval to disruptions in supply chains and field activities.

In Kenya, institutions such as the Kenya Medical Research Institute (KEMRI) and the Pharmacy and Poisons Board (PPB) of Kenya implemented interim guidelines that prioritized COVID-19 studies, revised consent procedures, minimized physical interactions, and adapted protocol requirements [6]. These changes, while necessary, had unintended implications on non-pandemic clinical trials. Studies not directly addressing COVID-19 faced delays, resource reallocation, and field constraints that threatened their methodological rigor and ethical compliance [7].

This paper presents a retrospective documentary case analysis of a non-COVID-19 clinical trial conducted during the pandemic in western Kenya: namely, the PRObiotics and SYNbiotics to improve gut health and growth in infants in western Kenya (PROSYNK Trial). This randomized controlled trial, conducted from October 2020 to January 2024, investigated the efficacy of probiotics and synbiotics in improving gut health and reducing systemic inflammation in infants [8]. Despite its non-pandemic focus, the study was shaped profoundly by the pandemic context. The documentary analysis reported here aims to highlight the ethical challenges and trade-offs encountered, as well as the adaptive strategies employed to maintain research quality, participant safety, and ethical standards in a dynamic and constrained environment.

## 2.0 Materials and methods

### 2.1 Study setting

The methods and results of the PROSYNK study have been published previously [8]. Only those aspects of the clinical trial relevant to the documentary analysis are reported here. Six hundred newborns under four days old were enrolled from Homa Bay County Teaching and Referral Hospital (HBCTRH) and nearby delivery centres in western Kenya. The infants were randomly assigned, with stratification based on HIV exposure, to one of four study groups in equal proportions (1:1:1:1) to receive either one of two synbiotics, a probiotic, or no supplement. Participants were scheduled to receive 32 doses of the assigned intervention over six months, administered through directly observed therapy (DOT) during home visits. The assigned supplements were administered daily for the first 10 days, then weekly up to six months of age. The participants were monitored until they reached two years of age. The primary outcome measure was systemic inflammation at six months, evaluated using plasma alpha-1-acid glycoprotein levels.

HBCTRH is located in Homa Bay town, the administrative and commercial centre of Homa Bay County in western Kenya. The hospital serves as a key referral centre for maternal and child health services in a region with high infant morbidity and limited healthcare access. The geographical setting, characterized by rural catchments and socio-economic vulnerabilities, was critical in shaping the study’s design and response during the COVID-19 pandemic.

### 2.2 Study design

The study reported here employed a retrospective documentary case analysis and reflective reporting methodology to explore the operational, ethical, and logistical experiences of conducting a non-COVID-19 clinical trial during a global pandemic. The approach enabled in-depth examination of the adaptations made in the midst of evolving regulatory, health system, and community-level challenges.

### 2.3 Case Selection

The PROSYNK trial was purposefully selected due to the first author’s direct involvement in its implementation. This involvement spanned regulatory submissions, community engagement, participant recruitment, follow-up procedures, and dissemination of findings. The PROSYNK trial was conducted amid heightened pandemic constraints and thus provided a suitable case for examining resilience and innovation in research conduct under crisis conditions.

### 2.4 Data collection and analysis

Archived PROSYNK study documents were retrieved from a central archival area at Kenya Medical Research Institute Centre for Global Health Research (KEMRI-CGHR) at Kisian, Kisumu. Key materials retrieved and reviewed included ethics and regulatory submissions, study Gantt charts, meeting minutes, progress reports, and sponsor communications. A chronological timeline of events was constructed and compared against the original project plan to map deviations and identify delays.

Additional data on the broader pandemic context such as health worker strikes and containment measures were gathered from peer-reviewed publications and mainstream news sources. All documents relevant to the research questions were logged systematically using a Document Access Log.

Thematic analysis followed a structured framework, focusing on regulatory delays, healthcare disruptions, and adaptive strategies. Insightful data were extracted, categorized, and refined iteratively. Emergent patterns were verified across multiple data sources to strengthen interpretive validity.

### 2.5 Reliability and validity

To ensure analytical rigor and consistency in data interpretation, systematic documents review was employed. Two trained research assistants independently examined all documents using a pre-defined, standardized coding framework designed to capture relevant themes, patterns, and categories systematically. This dual-review approach aimed to minimize individual bias and enhance the reliability of the coding process.

To assess the level of agreement between the two reviewers, Cohen’s Kappa statistic was calculated. This statistical measure accounts for agreement occurring by chance and provides a more robust evaluation of inter-rater reliability. A Kappa value greater than 0.6 was considered indicative of an acceptable level of agreement, reflecting substantial consistency between coders.

In instances where discrepancies arose between the reviewers, the documents in question were flagged for further analysis. The principal investigator then conducted an independent review of the contested items. Following this, the research team held structured debriefing discussions to resolve differences. Through these discussions, consensus was reached, ensuring that the final coding accurately reflected a shared interpretation of the data. This iterative process contributed to the methodological rigor and reliability of the study’s qualitative analysis.

### 2.6 Ethical considerations

The original PROSYNK trial (reported in [8]) received ethics approval from the KEMRI Scientific and Ethics Review Unit (SERU) on 16 October 2019 (KEMRI/SERU/CGHR/320/3917) and the Kenyan PPB on 5 August 2020 (ECCT/20/04/02). The Liverpool School of Tropical Medicine (LSTM) agreed to act as study Sponsor on 7 August 2020 (ref 19-048). To conduct the retrospective analysis reported here, an amendment was submitted to SERU and approved on 24 March 2025. A Data Sharing Agreement to accessed archived PROSYNK records was signed between the first author and LSTM on 26 January 2025. Only anonymized archived data were accessed on 31 March 2025. No participant-identifiable information was accessed, and all data handling adhered to established ethical and confidentiality standards.

## 3.0 Results

### 3.1 Regulatory timelines and study implementation

The PROSYNK study initially anticipated obtaining ethics and regulatory approvals within six months of submission, with participant follow-up projected to conclude by June 2023, marking 48 months from the study’s launch (Fig 1). The initial submission for ethics review was made on 17 June 2019, and approval was granted on 16 October 2019. However, a change in the investigational product required a protocol amendment, which delayed submission to the regulatory authority until 20 April 2020. Final regulatory approval was subsequently granted on 5 August 2020. To expedite the process, all protocol amendment submissions were made in parallel to both the ERC and the Regulatory Authority (RA), significantly reducing the overall time to approval.

**Fig 1.**
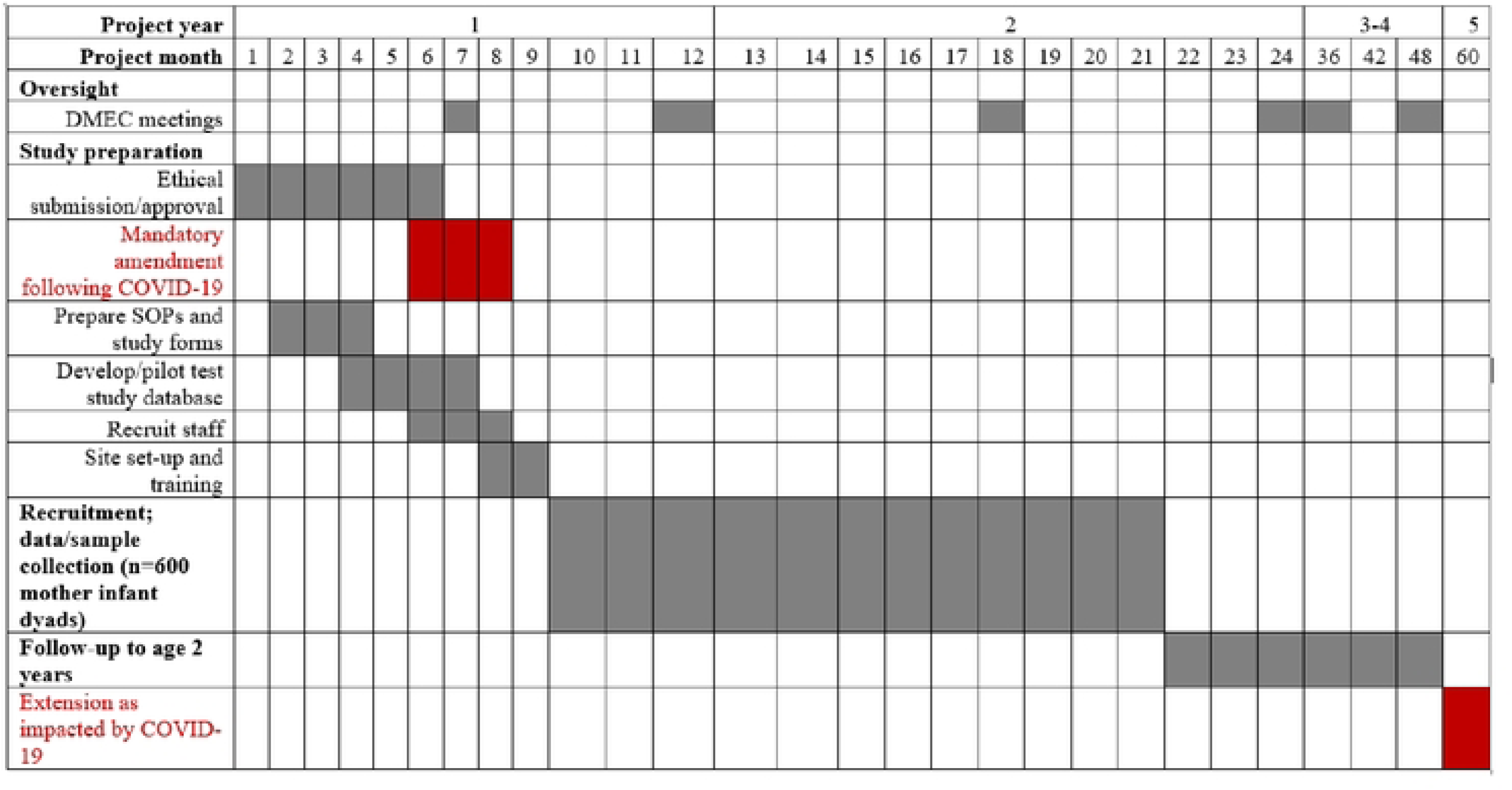
PROSYNK Study Gantt Chart as impacted by COVID-19.

Participant enrolment commenced on 28 October 2020, and final data collection concluded on 18 January 2024 representing a seven-month delay compared to the original timeline. The most substantial disruption occurred during Kenya’s third wave of COVID-19 (April–July 2021). During this period, the study sponsor, LSTM, issued enhanced safety guidelines via a memo dated 21 June 2021. These measures were consistent with the national Ministry of Health (MoH) COVID-19 protocols.

### 3.2 Recruitment and participant follow-up

HBCTRH, the primary recruitment site for the trial, adopted more stringent COVID-19 containment protocols than surrounding facilities. Operational challenges intensified during the third wave of COVID-19 in Kenya (April–July 2021), with mobility restrictions, infection control mandates, and periodic closures of hospital departments following staff exposure (Fig 2). HBCTRH temporarily closed due to confirmed COVID-19 cases among staff. With a shift in deliveries to peripheral sites and an increase in births attended by Traditional Birth Attendants (TBAs), participant recruitment was adversely impacted. Weekly reports submitted to the sponsor reflected a consistent shortfall in recruitment targets.

**Fig 2.**
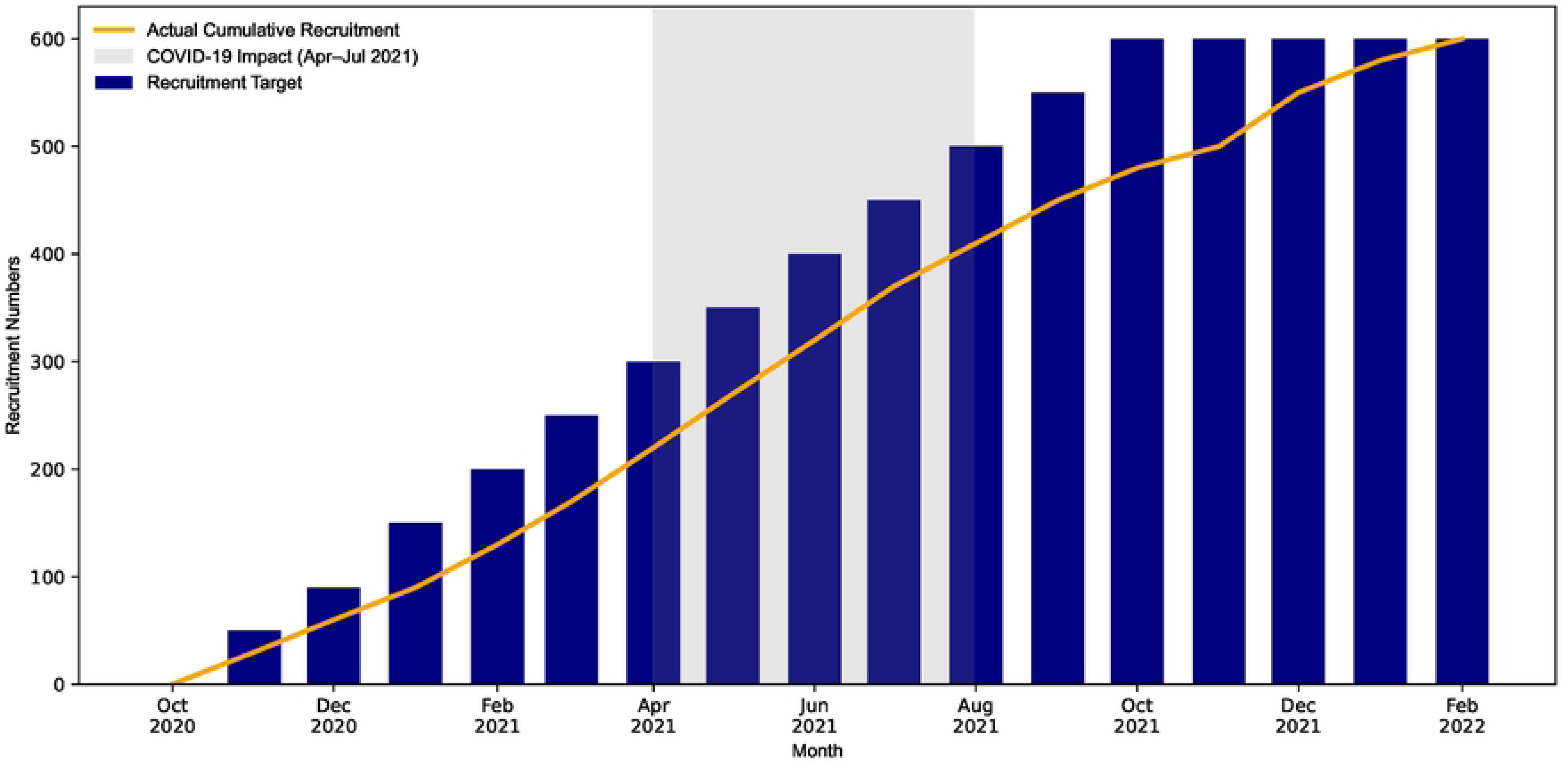
Monthly Recruitment vs. Target as impacted by COVID-19 containment measures.

Despite recruitment challenges, the study achieved a high follow-up completion rate: 546 out of 600 participants (91%) completed the study (Fig 3). Of the 19,200 planned home visits, those in the control group were substituted with phone calls. Despite this adjustment, a total of 18,412 visits/calls representing 96% of the planned interactions were successfully completed. Overall, the proportion of completed visits/calls during the COVID-19 3^rd^ wave was similar to that for the non-COVID-19 period. These visits, whether in-person or by phone, were used to administer supplements and/or deliver health messaging across both the intervention and control groups.

**Fig 3.**
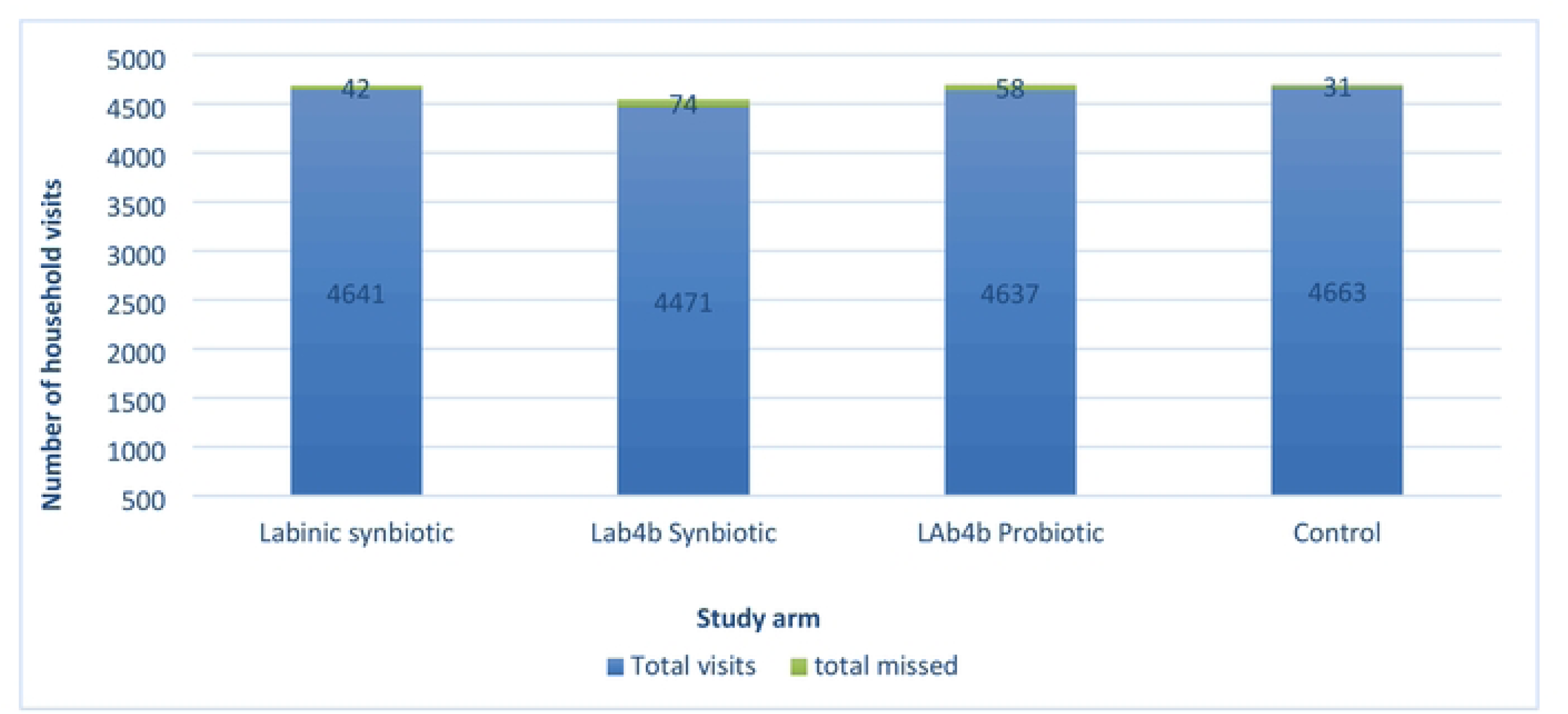
Supplement administration and missed visits by study arm.

### 3.3 Healthcare and socioeconomic impacts of the pandemic

COVID-19 containment policies significantly restricted access to healthcare facilities, complicating study recruitment and follow-up. Peer-reviewed studies and media reports documented nationwide health system strain, high infection risk among healthcare workers, and resulting industrial action. These factors, combined with localized COVID-19 outbreaks, led to intermittent departmental closures at the main recruitment centre, as noted in a news article dated 8 June 2021. Such disruptions underscored the volatile research environment during the pandemic period.

### 3.4 Adaptive strategies employed by the study team

To mitigate pandemic-related disruptions and ensure continuity of study, the PROSYNK team implemented a series of adaptive measures, documented in IRB submissions, meeting minutes, and study reports. These strategies aimed to protect participants and staff while maintaining methodological and ethical rigor. The initial study protocol gave room for the addition of satellite facilities to help drive recruitment through deployment of outreach teams. Introduction letters and flyers were circulated to the peripheral facilities to facilitate referral of mothers due for delivery to the trial facility.

The minutes of frequent study leadership meetings documented evaluation of prevailing COVID- 19 conditions over time. Recommendations were shared with the Data Safety and Monitoring Board (DSMB). The board advised on the course of the study following careful evaluation of risks to participants and staff.

Consultative meetings were held with community focal persons and enlisted TBAs, resulting in an agreement to support appropriate referrals of expectant mothers to the trial facility. The study team also engaged an active Community Advisory Board (CAB) to facilitate ongoing community dialogue. Minutes from these meetings highlighted the team’s efforts to dispel myths surrounding COVID-19 and to communicate additional safety measures implemented to protect both participants and the wider community.

On 31 July 2020 a mandatory amendment was submitted to the ERC in accordance with updated COVID-19 guidelines for research studies. The amendment was for the use of phone calls, in place of home visits, for participants in the control arm where supervised dosing was not required.

Clear communication was maintained through internal memos outlining the specific measures to be implemented due to COVID-19. To adapt to the evolving situation, the study team adjusted the enrollment target to a more manageable figure reducing it from twelve to six participants per week. Additionally, research staff operated in rotating teams, a strategy that helped minimize crowding at the research center. The work teams were balanced with each comprised of study team members able to conduct all study procedures adequately. The study coordinator ensured smooth transitions between outgoing and incoming teams by prioritizing all pending work assignments.

In addition to prioritizing the standard of care for all participants, the study team facilitated COVID-19 testing for individuals presenting with symptoms of severe acute respiratory illness. As the host institution’s laboratory was an approved COVID-19 testing center, the study team was able to ensure a rapid turnaround of results to support timely clinical decision-making and patient care. Additionally, Personal Protective Equipment (PPE) was provided to general hospital staff and remedial measures were undertaken to curb the spread of COVID-19, including routine COVID-19 testing for study staff during work cohorts and disinfection of workspaces.

Study tasks and activities were carefully sequenced, with timelines for procedures such as visit windows considered to ensure smooth implementation. Communication channels were established to support effective collaboration and timely sharing of information among team members. The CommCare App was utilized to monitor scheduled clinic visits and track when supplement doses were due for participants.

## 4.0 Discussion

The PROSYNK study provides valuable insights into the conduct of non-COVID-19 clinical research during a global health crisis. Navigating a shifting regulatory landscape and operational uncertainty, the study highlights the challenges and opportunities in maintaining research continuity during a pandemic.

### 4.1 Operational disruption

The PROSYNK study experienced significant delays that extended its planned closure date from June 2023 to January 2024. These delays disrupted the overall study timeline and reflected broader global patterns of regulatory slowdown during the COVID-19 pandemic. Ethics review boards and regulatory authorities were widely reported to have reallocated attention toward COVID-19-related research priorities [9,10].

Operational disruptions were most acute during Kenya’s third wave of COVID-19 (April–July 2021), when government-imposed mobility restrictions, infection control protocols, and intermittent hospital closures directly impacted research activities. For example, on 8 June 2021, entire departments at HBCTRH, the study’s primary recruitment site, were temporarily shut down following confirmed staff infections [11]. These events echoed national trends in healthcare system strain, including labour unrest and a resurgence of traditional birth practices, as documented in Coastal Kenya [12].

Further, healthcare workers’ strikes in Kenya during the COVID-19 pandemic significantly compounded the public health crisis. Most medical personnel in public hospitals engaged in industrial action, disrupting essential health services at a critical time. The strikes were driven by demands for adequate insurance coverage and sufficient personal protective equipment (PPE), both of which were lacking. These protests occurred amid rising COVID-19 cases and mounting pressure on the health system. The situation was further exacerbated by the deaths of at least 14 doctors from COVID-19 since the pandemic began, underscored the urgent need for improved occupational safety and support for frontline health workers [13].

Within this context, achieving recruitment targets became increasingly unfeasible. These challenges highlight the fragility of facility-based studies during public health emergencies and underscore the need for adaptive research designs that can withstand future operational disruptions.

### 4.2 Adaptive and ethical research management

Despite the considerable operational challenges faced by the PROSYNK study, the research team exhibited exemplary ethical leadership through resilience and adaptability. Key mitigation strategies were implemented to uphold the study’s integrity and participant welfare, including expanding recruitment efforts to satellite health centres, collaborating closely with community focal persons and traditional birth attendants (TBAs), rotating staff teams to reduce COVID-19 exposure risk, and strengthening infection prevention protocols. These measures reflect core ethical principles of beneficence, justice, and respect for persons [14] ensuring that participant safety and equitable access remained paramount even amid a public health crisis.

Central to this ethical leadership was the deliberate focus on community trust-building. The team maintained frequent, transparent communication with healthcare workers, community stakeholders, and oversight bodies such as the study sponsor. This openness helped counter misinformation, foster accountability, and support informed, ethically sound decision-making throughout the study. As Mwangi et al. highlight, such strong leadership combined with inclusive dialogue is vital for sustaining research momentum and ethical rigor in times of uncertainty [15].

This approach underscores how ethical leadership can effectively navigate complex challenges, balancing scientific goals with the moral imperative to protect and respect participants and communities during crises.

### 4.3 High follow-up and participant retention

The study notably achieved a 91% follow-up completion rate and a 96% success rate in scheduled home visits/calls, demonstrating the commitment and resilience of the field teams. A key factor contributing to these outcomes was the integration of remote follow-up options for the control group, alongside the use of digital platforms such as CommCare. These technological adaptations facilitated uninterrupted data collection while reducing health risks associated with in-person contact. Such innovations highlight the critical role of flexible research designs and digital tools in maintaining data integrity and study continuity amid operational constraints, aligning with broader evidence on the benefits of digital health technologies in research during public health emergencies [16,17].

Priority home visits were conducted for participants who had been scheduled for phone follow-up but could not be reached. Consistent with findings by Houlding et al., limited access to mobile phones was observed among participants. These challenges highlight the need for developing a structured framework for remote monitoring in research, particularly in low-resource settings [18].

The implementation of COVID-19 control measures in Kenya varied notably across regions, shaped by local epidemiological trends, socio-economic conditions, and administrative frameworks. In rural areas such as the setting for this study, the enforcement of these measures was relatively less stringent, which allowed for the continued progression of research activities with minimal disruption [19].

### 4.4 Ethical responsiveness in public health emergencies

The study team demonstrated a proactive and ethically grounded response to emerging public health threats, going beyond protocol modifications. The provision of COVID-19 testing for symptomatic participants and staff, as well as routine testing, provision of PPE, and regular disinfection of workspaces, underscore a strong commitment to the safety and wellbeing of both research personnel and community participants, an often underreported but critical aspect of conducting research during public health crises. As Moodley K. et al. suggest, leadership in healthcare and research must proactively address systemic barriers to both service delivery and study participation during emergencies [20].

## 5.0 Conclusion and recommendations

The PROSYNK study illustrates both the vulnerability and resilience of clinical research in the face of global health emergencies. While delays caused by public health restrictions disrupted the original study timeline, the research team’s adaptive strategies grounded in ethical responsiveness, community engagement, and operational flexibility enabled the study to meet its core objectives. Recruitment to target, high participant retention rates with follow-up to study completion, despite significant constraints, affirm the effectiveness of decentralized recruitment models, strong stakeholder collaboration, and technological innovations.

These findings highlight critical lessons for sustaining research in dynamic, resource-limited, or crisis-affected settings. Researchers, ethics bodies, and sponsors alike must be prepared to adjust traditional processes to uphold ethical standards while maintaining scientific rigor. The PROSYNK experience underscores the value of proactive, community-centric, and flexible research design, particularly when navigating the uncertainties of public health emergencies.

To strengthen future research implementation during pandemics or similar disruptions, the following recommendations are proposed:

1. Regulatory Flexibility with Ethical Vigilance Ethics review committees should consider allowing the continuation of non-pandemic research where clear benefits exist and risks to participants, staff, and the broader community are demonstrably mitigated. A nuanced, risk-benefit approach is essential to avoid blanket suspensions that may inadvertently harm vulnerable populations.
2. Transparent Stakeholder Engagement Researchers should foster regular, open communication with all stakeholders including participants, community leaders, sponsors, and oversight bodies to promote trust, reduce misinformation, and support ethical decision-making under evolving conditions.
3. Proactive Risk Monitoring and Response Research teams must remain responsive to emerging threats by monitoring local trends and implementing context-appropriate mitigation strategies. Ensuring participant and staff safety through timely adjustments—such as remote follow-ups or PPE provision—can preserve study integrity while minimizing harm.
4. Flexible and Decentralized Research Models Incorporating contingency plans into study protocols enhances resilience. The successful use of satellite sites, community health workers, and TBAs in PROSYNK demonstrates the value of decentralized recruitment strategies. Embedding such models at the design stage can improve reach and retention, especially in underserved or disrupted areas.

These recommendations advocate for a more agile approach to research in complex environments that remain ethically grounded. As the global research community continues to contend with uncertainty and rapid change, the lessons from PROSYNK serve as a roadmap for future preparedness, continuity, and ethical excellence in clinical research.

## Data Availability

Data supporting the findings of this study are available upon reasonable request. Interested parties should contact the corresponding author via email. Access will be granted following approval by all principal investigators involved in the original study.

## Acknowledgments

The authors gratefully acknowledge the contributions of all PROSYNK trial participants and the dedicated staff at the Kenya Medical Research Institute (KEMRI)/Centre for Global Health Research in Kisumu, Kenya, for their efforts in data collection and trial implementation.

## Supporting information

**S1 Fig 1. PROSYNK Study Gantt Chart as impacted by COVID-19**

**S2 Fig 2. Monthly Recruitment vs. Target as impacted by COVID-19 containment measures**

**S3 Fig 3. Supplement administration and missed visits by study arm**

**S1 File 1. Supplementary Table 1. Protocol amendment and approval timeline**

**S2 File 2. Supplementary Table 2. Framework analysis of key themes influencing PROSYNK Trial conduct during the COVID-19 Pandemic**

**S3 File 3. Supplementary Fig 1. Original PROSYNK Study Gantt Chart (2 years follow-up to 48 months)**

**S4 File 4. Supplementary Fig 2. Conceptual framework**

